# New WHO guideline on the definition of anemia: implications for 6-35 months old children in Peru 2009-2023

**DOI:** 10.1101/2024.05.28.24308069

**Authors:** Miguel Campos-Sánchez, Luis Cordero Muñoz, Enrique Velásquez Hurtado, Nelly Baiocchi Ureta, Marianella Miranda-Cuadros, María Inés Sánchez-Griñán, Walter Valdivia Miranda

## Abstract

**Introduction:** The World Health Organization recently published a guideline updating the cut-off points defining anemia, modifying the age and altitude adjustments and recommending auto-analyzers. We compute in a national sample the magnitude of anemia prevalence with the previous and current guidelines between 6 and 35 months old and discuss the implications.

**Methods:** Secondary analysis of the Peruvian Demographic and Health Survey 2009-2023, a repeated cross-sectional design upon a national stratified and cluster random sample.

**Results:** 117995 children were included. With the new guideline, the national prevalence is reduced and the regional prevalences are generally reduced (some increased) in variable amounts.

**Conclusions and Recommendations:** The comparison confirms that the new guideline modifies, mostly reducing, the prevalence in an important and heterogeneous magnitude. Literature supports the direction, but not the magnitude of the correction. We recommend the quick, but cautious and gradual adoption of the new guideline. For analytic calculations, age and altitude formulas (not categories) should be used. Surveillance must be reinforced and control strategies must be reviewed. Case management guidelines must be updated.

## Introduction

Children anemia is a major national and international public health problem (1–5) particularly by its consequences on early development. The World Health Organization (WHO) published last 06-Mar a new guideline (6) on the hemoglobin cutoff points which define anemia, updating a previous guideline (7,8) taking into account important and recently published evidence (9–11). The new guideline has three main changes for small children: it reduces the cutoff point for the 6-23 months old group, it modifies the altitude adjustment equation (lowering the cutoff points for altitudes above 3000m, while rising the cutoff points for altitudes below that level) and it requires venous blood measurement using auto-analyzers or calibrated portable devices, both under quality control. Peru has updated its technical norm (12) with the new guideline.

In the present paper we carry out a comparative analysis of the prevalence of anemia using both guidelines and we discuss the implications for the health status of the population of children between 6 and 35 months old residing in Peru between 2009 and 2023.

## Methods

This paper carries out a secondary analysis of data from the Peruvian Demographic and Health Survey (ENDES) (13,14) from 2009 to 2023, carried out by the Instituto Nacional de Estadística e Informática (INEI), since 1986 as a DHS surveys, continuous since 2004.

ENDES has a repeated cross-sectional design upon a random national sample. The sampling frame, mantained by INEI, divides the whole country in clusters having approximately 120-140 households each, distributed among strata by first level administrative regions and socioeconomic level, within each year. INEI computes the sampling size as a multipurpose survey, having approximately 35000 households each year. In the first stage, clusters are selected from each stratum with probability proportional to size and distributed along the calendar months. In the second stage, the list of households in each cluster is updated by the field teams and a random sample is selected for interview. Age (birth date) and sex are obtained by declaration. Rural towns are defined if they have less than 2000 inhabitants. Altitude (m above sea level) is defined by INEI for each cluster (since 2016 INEI gets GPS household altitudes, a datum not used here). In subsamples of several population subgroups hemoglobin is measured on capillary blood using portable hemoglobinometers (personnel is standardized at least once a year). INEI continuously updates the sampling frame, readjusting the rural classification of clusters and the sampling techniques every now and then. Anonymized individual data as well as documentation is publicly available.

Individual data were read and consolidated, readjusting sampling weights for each year according to the national population projections (15) and redefining strata as 25 regions x 2 settings (urban and rural) each year. Children were included in the analysis if they had between 6 and 35 months old and had non missing hemoglobin reading, for a total of 117995 children. Estimation of prevalence proportions was carried out for each guideline (6,8) with weighing adjusted for the sampling design (16,17). R software (18) version 4.3.1 with tidyverse (19), and survey (20) packages and their dependencies were used. Code and consolidated data for this paper have been made publicly available as file CUTHBN32024.zip at https://github.com/vipermcs/pdata/.

## Results

Figure 1 shows the national prevalence trend for each guideline.

**Figure 1.**
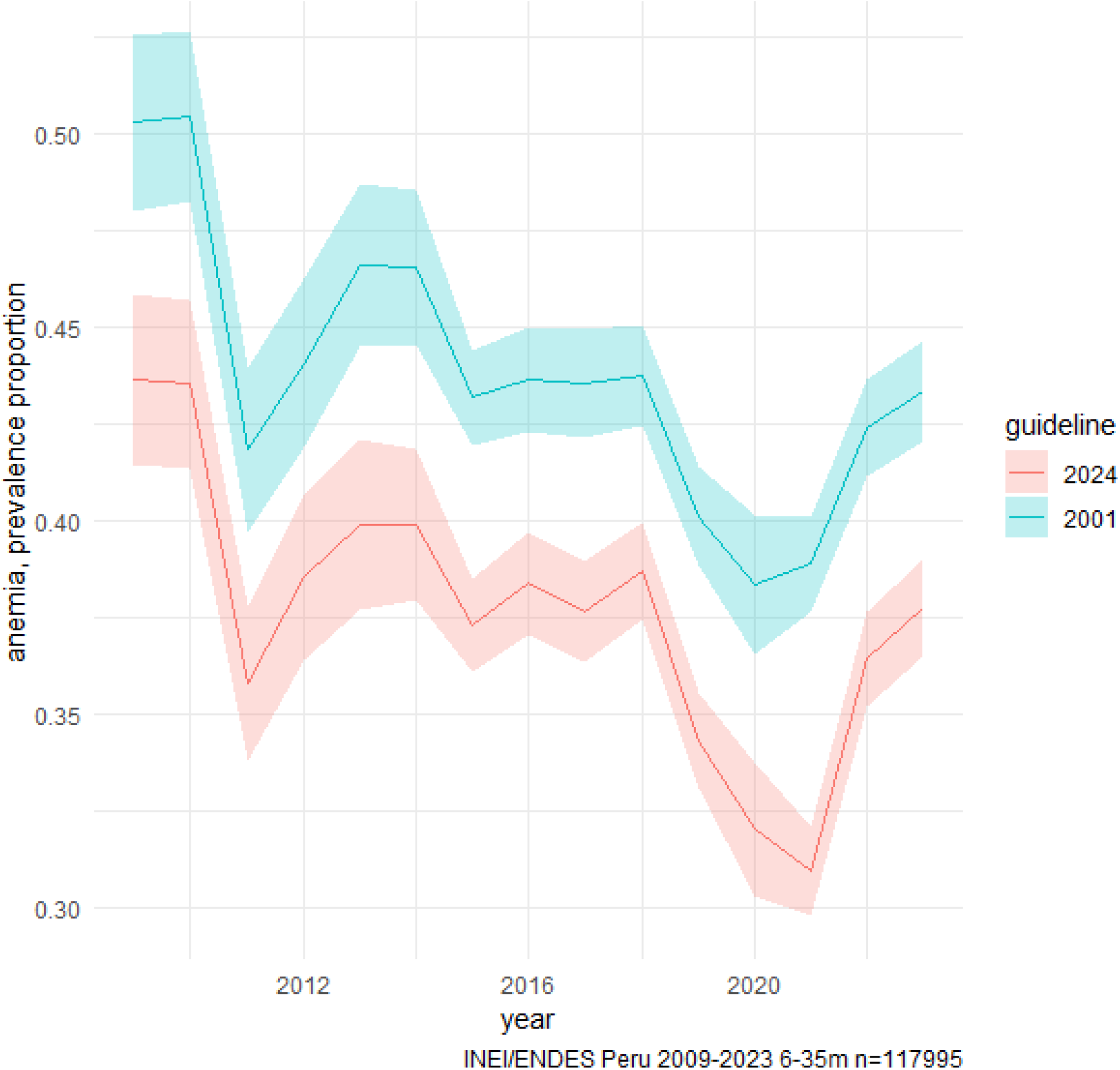
Trends in National Prevalence

Lines join yearly estimates. Bands mark their 95% confidence intervals.

We can see that the prevalence pattern over time is similar for both guidelines (within the confidence limits). With the new guideline the prevalence is clearly lower than the old and below the 0.40 level which officially (21) divides severe and moderate public health problems, deciding the recommendation of universal daily supplementation in the range 6-23m.

Figure 2 shows the prevalence trend over time for both guidelines at each combination of setting, age group and grouped altitude level.

**Figure 2.**
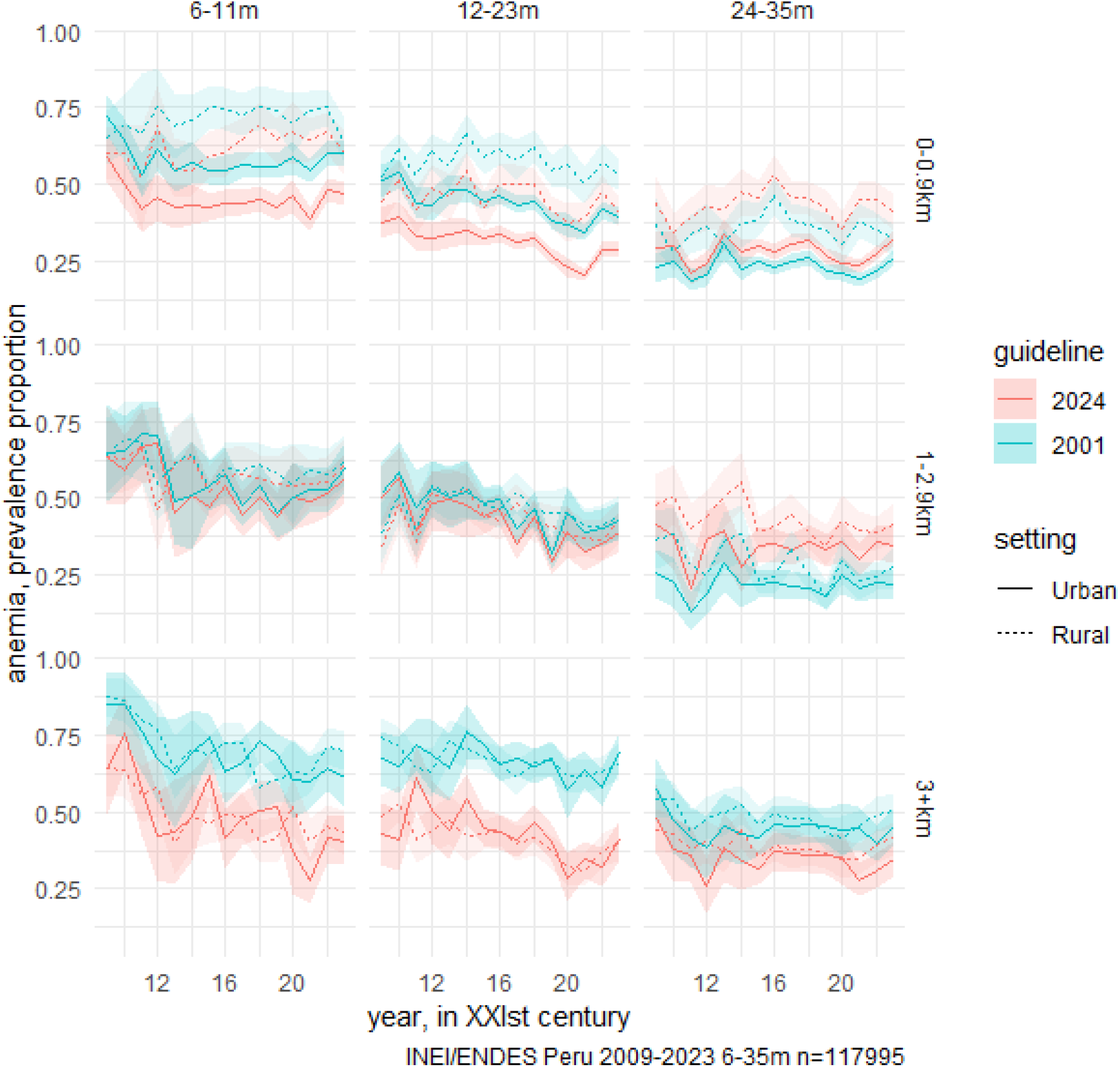
Trends in Subnational Prevalence

Lines join yearly estimates. Bands mark their 95% confidence intervals.

We can see that, among children 24 months old or more (for whom the cutoff points in both guidelines are the same) differences in the estimated prevalence of anemia appear, generated by the change in the adjustment equation for altitude. Such differences are non-linear, showing and increase in the prevalence of anemia when the altitude is below 3000 m. In children under 24 months old, the difference in the prevalence of anemia between both guidelines originates from both the change in the adjustment equation for altitude and the reduction in the sea level cutoff point in this age group.

Figure 3 presents the prevalence of anemia comparing both guidelines for each of the 25 first level administrative regions and each of the 14 years.

**Figure 3.**
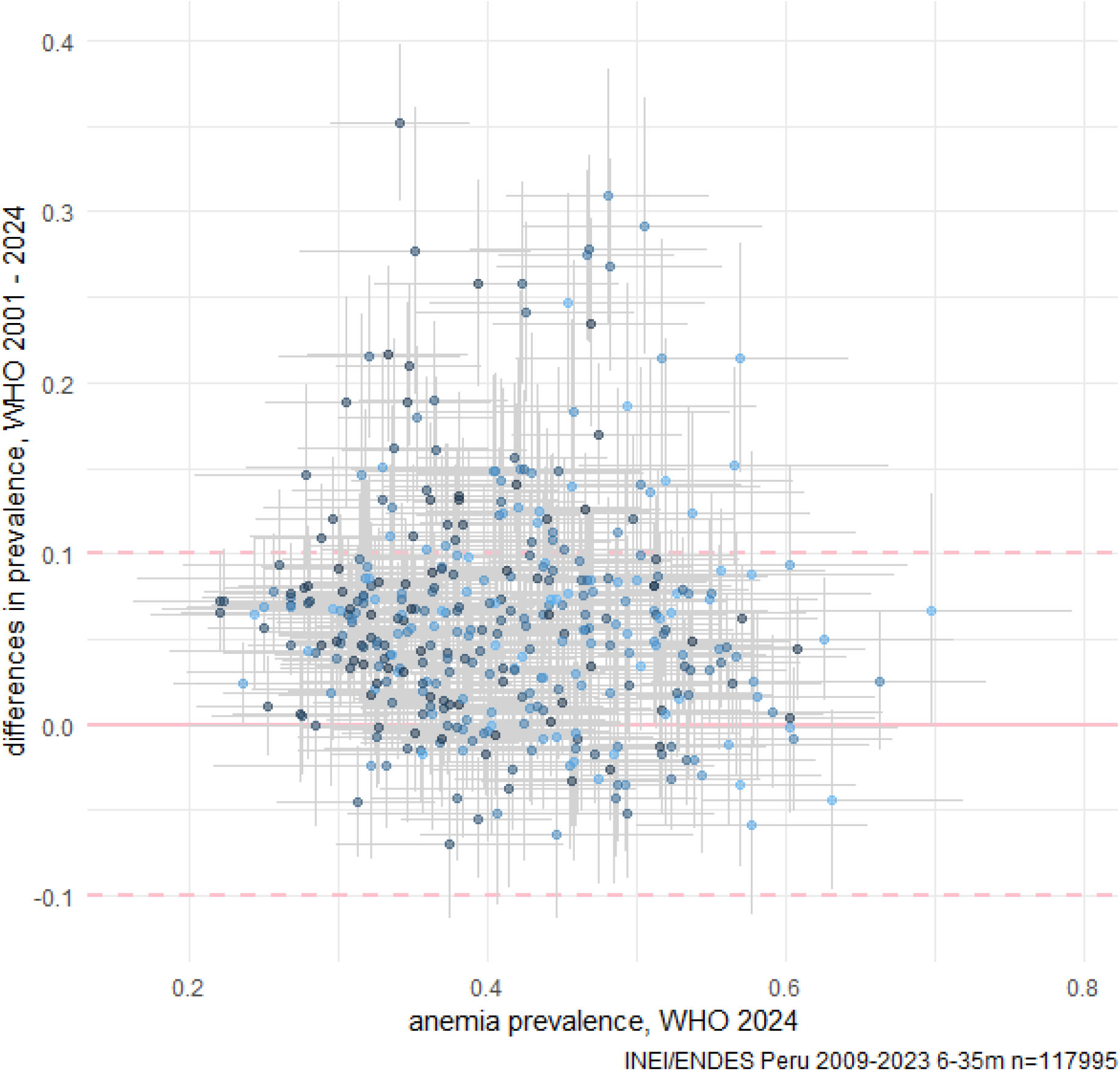
Prevalence by Region-Year, comparing guidelines

Each point is a region in a year. Colors are lighter in early years, toward 2009, and darker toward 2023. The pink line is the identity. Gray segments are the 95% confidence intervals.

We can see that there is no simple correspondence between the prevalences of anemia calculated with both guidelines. Several points reduce their level, while some increase theirs.

The guidelines describe two ways to adjust for altitude: a classification table and a formula. The table does not define an adjustment for 5000 m or more. The 2001 guideline specified that below 1000 m no adjustment was applied. The formula is a continuous curve, the table is a staircase line. Figure 4 compares prevalences for each region-year computed both ways.

**Figure 4.**
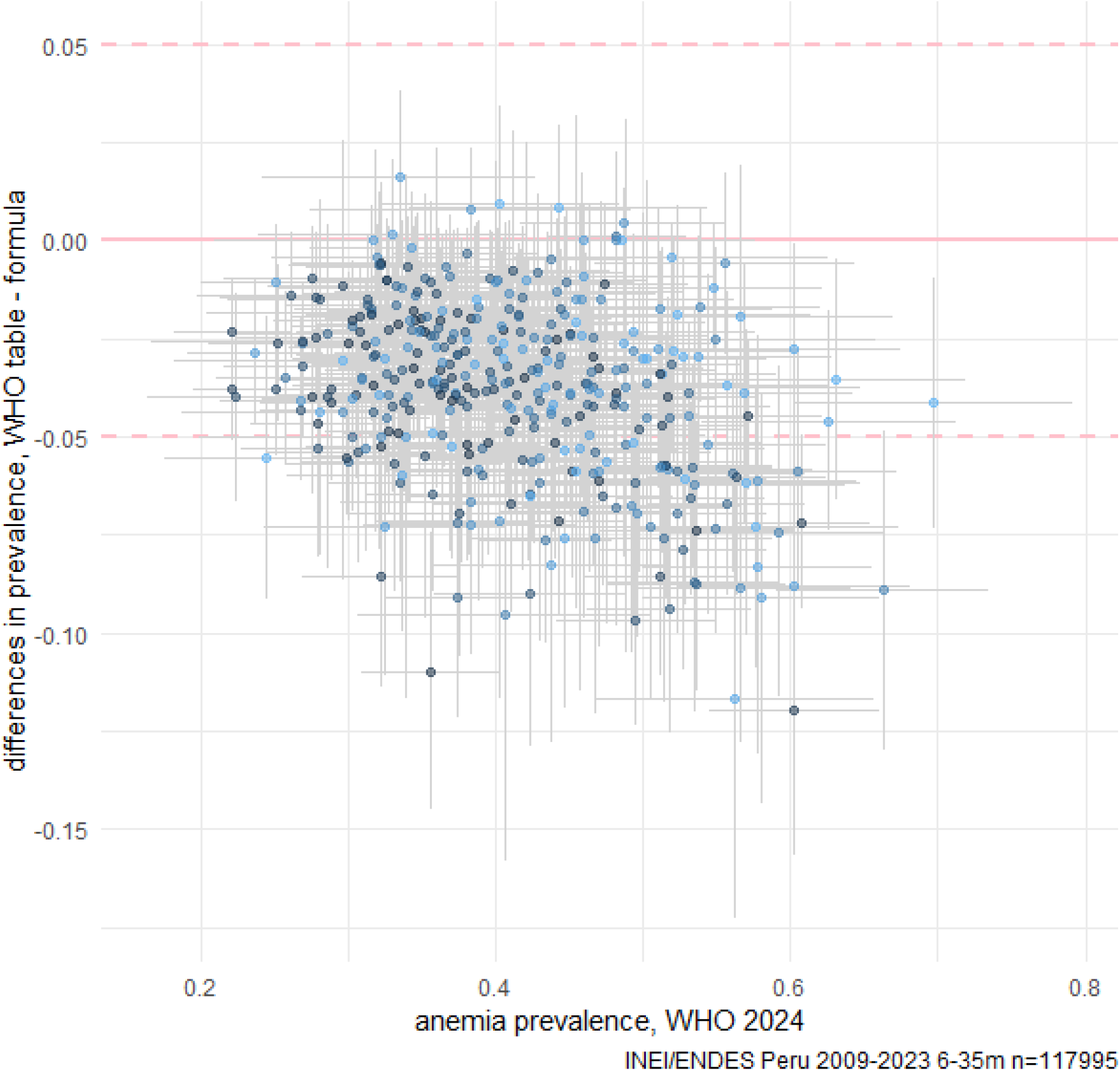
Prevalence by Region-Year, comparing technique

Each point is a region in a year. Colors are lighter in early years, toward 2009, and darker toward 2023. The pink line is the identity. Dashed lines mark differences in prevalence of 0.05 around the identity. Gray segments are 95% confidence intervals.

We can see that the prevalence of anemia is markedly different for both ways. In general, but not always, the table underestimates the prevalence, reaching differences of 0.05 and even 0.10. In this paper we applied the formula for all altitudes from 0 m upwards.

## Discussion

With the new guideline our analysis finds a reduction in the national prevalence and a change (mainly, but not always, a reduction) in the age and altitude subgroup prevalences. The time patterns are similar, but not identical, with both guidelines. This arises from the non-linearity of the guideline changes, the population distribution by altitude and the hemoglobin distributions being near the cutoff points.

We can identify the following limitations: (a) the re-scaling of sampling weights, which we do not think it distorts the calculations, which are by year and do not apply modeling; (b) the changes in sampling design (14,22) at ENDES trienal cycles, such as the definition of strata, which we redefined (we cannot exclude with certainty changes associated to the design modifications); (c) the precision, particularly for subgroups with smaller samples, so small differences cannot be surely established; (d) the measurement on capillary blood with portable HemoCue (mostly model 201) devices; (e) the definition of a single altitude for each cluster, which may have households at very different altitudes.

On the other hand, the ENDES data source has important strengths: national and subnational representativeness, its continuity for more than a decade, and the ecologic, economic, social and cultural diversity of Peru.

It is logical that the new guidelines generate changes in the prevalences. What our analysis adds is an appreciation of the magnitude of those changes in a real and diverse population. We will review the literature review and reflect around two issues: the support for the new guideline and the advisability of its adoption.

### First issue ¿Is the 2024 guideline correct?

We shall review if the existing evidence is enough to support the statement that the new guideline corresponds to biologically optimal hemoglobin values by age and altitude.

The 2001 guideline (7,8) lacked solid evidence, being essentially an expert opinion quoting a few studies. There were hints that the 1968 and 2001 guidelines overestimated the cutoff points for the first months of life (23) and the altitude adjustments (11), likely nproducing a overestimation of the prevalence of anemia.

A wide international (23–25) and national (26–29) debate arose. Our interpretation of the consensus up to 2023 is to acknowledge that the evidence supported the need to modify the guidelines, as well as the direction in which the adjustments had to be done, but it did not support the magnitude of the necessary corrections.

The estimation of normal values of hemoglobin by age and altitude ideally requires cohort studies on representative samples of large populations, as well as complementary studies. The new guideline instead uses what it calls “statistical approach” (common in clinical pathology). That is the estimation of distribution percentiles from national representative surveys (and some non representative studies) with variables (iron and inflammation markers and symptoms) which allow to select a “healthy” subset. This approach could crucially fail when the prevalence of some causes of anemia is not small and diagnostic methods to exclude them are not available.

The new cutoff points are based on the following evidence (which represents somewhat less than 11% of the world population):

- One paper on age (9). Its relevant data sources (1999-2019) are two surveys (USA and Ecuador), and a cohort (Canada/TARGet Kids!). Its analysis fits a curve over the second semester of life. This better reflects the physiology than the abrupt transition in the guideline, which generates an artificial jump at 24 months of age.
- Two papers on altitude for children <60 months old and women 15-49 years old (11) and school age children (10). Their relevant data sources (1999-2019, BRINDA project) are surveys (Mexico, Colombia, Ecuador, Afghanistan, Nepal, Laos, Malawi, Papua New Guinea, Azerbaijan, Georgia, UK and USA), a surveillance (Guatemala/SIVESNU), and two evaluations (Ghana/GIFTS and Bolivia/NIDI (El Alto)). Besides, the 2024 guideline describes an extension (“Pompano, CDC, unpublished data, December 2023”), adding 5 years of the Peru/SIEN surveillance (the only source above 4000 m). In children <60 months old (11) some between country heterogeneity is shown. The equation in the 2024 guideline is nearly identical to the school age fit (10).

We can add the following evidence:

- Two studies mentioned by the 2024 guideline as having different inclusion criteria. One is a BRINDA review (30) with precursor evidence examining cutoff points for the 6-59 months old pooled together finding wide heterogeneity and a departure from the 2001 guideline. The other is a survey from India (31) which, having intention and approach (although not method) similar to WHO, proposes cutoff points markedly below the 2001 and 2024 guidelines.
- Previous studies supporting the existence of discrepancies with the 2001 guideline by age (32–38,23) and altitude (39,40), as well as the heterogeneity of such discrepancies (41,11,23), but not their magnitude. Of particular interest are some hints that the altitude adjustment may vary with evolutive adaption (42).
- A recent Peruvian study (43), whose sample (6-8 months old at sea level) is very small and whose representativeness is arguable (health center children without documentation of random selection). This study included, besides Lima, Arequipa, Cusco and Puno cities. Its exponential regression fit is different from the 2024 guideline. It seems likely that they will soon publish other age groups.
- Alternate adjustment proposals (44–47) whose support is very weak because of limited representativeness of their populations and limited ability to exclude pathology.

Our interpretation is that the current evidence, although strongly suggests that the 2024 guideline corrects in the proper direction, is not enough to prove its hemoglobin cutoff points are optimal and universal. The more important doubts are the representativeness of the reference populations, the inclusion and exclusion criteria, the smoothness of the age adjustment function and the heterogeneity between and within countries.

### Second issue ¿Must the new guideline be adopted by Peru?

Although it is extremely desirable to support decisions with scientifically solid evidence, in Public Health practice that is not always possible. It becomes necessary to make decisions under uncertainty, but reasonable precautions must be taken to opportunely manage strategic and tactical failures.

The hints of a need for correction are growing strong, but the definitive evidence on the magnitude of such correction may take years of research and most surely will produce additional modifications. The available evidence must be critically read and cautiously and gradually applied. The recommendations, in our opinion, are:

a. On the specific issue of the cutoff point:
  - To acknowledge the 2024 guideline as a provisional correction, not wholly supported.
  - In the analysis scenario, apply the formula, not the table, for altitude, starting at sea level; also apply the formula for age (9), even if not specified in the guideline. The computations must be interpreted as a mitigation with remaining limitations. The interpretation at middle and high altitude is fragile.
  - In the communication scenario, consider transition periods with parallel indicators, emphasis in transparency of methods and explanations.
b. On the scenario of Child Anemia control in Public Health:
  - Given the unresolved uncertainty, <u>reinforce</u> surveillance to detect low coverage, filtrations, and inequities (not just for hemoglobin indicators, but also their determinants, including social, environmental and interventions), as well as the consequences of anemia and its interventions (including side effects).
  - <u>Review</u> the strategy, reanalyzing the evidence, with some margin for intuitive decisions, so that, if that is the technical consensus, the target populations, the intervention profile, and the immediate research priorities are modified.
c. On the scenario of preventative and recuperative care of Individual Health:
  - Acknowledge that medical practice must apply its criterium to each unique patient.
  - <u>Update</u> individual management guidelines and algorithms (48–50).
  - <u>Review</u> the governmental plans for health services, insurance coverages, and health care procedures, particularly in Health, Social Programs and Social Security sectors.

The evidence gaps are challenges for short as well as long term research. Several studies which now are partial evidence could become baselines for cohorts to measure the risk of several outcomes as a function of hemoglobin, age, altitude and other covariables.

We have examined only the 6-35 months old subgroup. Similar considerations may affect other subgroups. We do not deal here with two also critical problems: etiology (51), particularly the real contribution of iron deficiency (52,53), and the instruments (54–57).

As a final reflection, we note that the 2024 guideline is conceptually retroactive. This logically implies that we, the technical community, were wrong in the individual and population diagnosis and subsequent management of anemia.

## Data Availability

All data and code in the present article has been made publicly available online as stated in the manuscript.

https://github.com/vipermcs/pdata/blob/main/CUTHBN32024.zip

## Notes

### Competing Interest Statement

The authors have declared no competing interest.

### Funding Statement

The study performed for this article did not receive any funding.

### Author Declarations

The study used only data that were originally located at https://www.datosabiertos.gob.pe/ and https://www.gob.pe/institucion/inei/tema/biblioteca-inei-8947dde8-6caf-4db9-a1be-397f79bbe65f as stated in the manuscript.

